# Effects of Health Technology Use and Digital Health Engagement on Clinical Trial Participation: Findings from the Health Information National Trends Survey

**DOI:** 10.1101/2024.09.13.24312295

**Authors:** Nicholas C. Peiper, Stephen Furmanek, Kelly McCants, Edward H. Brown

## Abstract

**Background:** Clinical trials are critical to scientific advances and medical progress, although awareness and participation remain low in the general population. The existing literature indicates that clinical trial knowledge and participation is multifactorial. Yet, little is known about the association between clinical trial participation with health technology use and digital health engagement to search for health information, interact with medical providers, and seek health supports.

**Objective:** Examine the multivariate association between clinical trial knowledge and participation with past-year health technology use and digital health engagement with medical providers.

**Design:** Cross-sectional data from a federal surveillance system.

**Participants:** A total of 3,865 US adult respondents from the Health Information National Trends Survey 5, Cycle 4 conducted in 2020.

**Main Measures:** The two outcomes were clinical trial knowledge (no knowledge, a little knowledge, a lot of knowledge) and participation (never invited, invited did not participate, invited and participated). There were four binary indicators of health technology use for the following purposes in the past year: searching for health or medical information, communicating with a doctor’s office, looking up medical test results, and making medical appointments. There were four binary indicators of digital health engagement in the past year: sharing health information on social media, participating in a health forum or support group, watching health-related videos on YouTube, and awareness of ClinicalTrials.gov.

**Key Results:** Survey-weighted multivariate regression models demonstrated that awareness of ClinicalTrials.gov had the largest associations with clinical trial knowledge and participation. Digital technology use to engage with medical providers and electronic health records was associated with clinical participation, although the vast majority of respondents had never been invited.

**Conclusions:** Findings from this study can inform the design of large-scale digital health campaigns and quality improvement programs focused on increasing clinical trial participation.

## Background

Clinical trials are critical to scientific advances and medical progress. As of July 2024, there are 501,869 registered studies on ClinicalTrials.gov, of which 67,295 are recruiting participants.^1^ These studies include a wide variety of scientific disciplines and include studies to test the efficacy of prevention, treatment, and screening interventions. Despite the benefits of clinical trials, they remain difficult to implement largely due to issues surrounding enrollment and retention such as transportation barriers, travel requirements, fear of adverse events, and medical distrust.^2–5^ Furthermore, adequate inclusion of women, racial and ethnic minorities, and other health disparity populations continues to be a large challenge.^6,7^ These findings illustrate a critical need to more thoroughly understand the factors that may improve clinical trial knowledge and participation.

The existing literature indicates that clinical trial knowledge and participation is multifactorial. For example, several national studies have recently found lower clinical trial knowledge and participation among certain demographic groups, including older adults, racial and ethnic minorities, people living in rural localities, and patients with medical comorbidities.^8–12^ Other studies have found that social determinants of health (SDOH) play an important role in clinical trial knowledge and participation.^7^ For example, Williams and colleagues (2023) found that financial barriers (e.g., lack of insurance coverage and psychosocial supports) to clinical trial participation were common (>50%) in a nationally representative sample of US adults.^13^ Yet, less is known about how health technology use and digital health engagement are associated with clinical trial knowledge and participation.

Understanding health technology use and digital health engagement is critical in identifying potential strategies to increase clinical trial knowledge and participation. Studies indicate that using digital technologies to search for health information and engage with healthcare providers is associated with higher health literacy and satisfaction with clinical services.^14–17^ Other studies have found low awareness of ClinicalTrials.gov and other digital health platforms in both clinical and population samples throughout the world.^18,19^ Because clinical trial awareness remains low in general populations throughout the world,^20^ studies that investigate the association between health technology use and digital health engagement with research participation can inform the design of large-scale digital health campaigns and quality improvement programs focused on increasing diversity in clinical research.^21^

To address this need, this study investigated the association between health technology use and digital health engagement with clinical trial knowledge and participation in the general population. The primary objective was to examine multivariate associations between clinical trial knowledge and multiple measures of health technology use and digital health engagement in the past year. The secondary objective was to examine multivariate associations between clinical trial participation and multiple measures of health technology use and digital health engagement in the past year. We hypothesized that health technology use and digital health engagement will be significantly associated with higher clinical trial knowledge and participation after adjusting for demographic and social factors.

## Methods

### Study Design

We used data from the Health Information National Trends Survey (HINTS), a federal surveillance system sponsored by the US National Cancer Institute, to collect nationally representative data about the American public’s use of health-related information.^22^ The HINTS program monitors various changes in the pattern of health information seeking and usage over time. This study focuses on the HINTS 5 Cycle 4 (H5C4) that collected data from non-institutionalized civilian adults ages 18 and older in 2020. HINTS applied a two-stage, stratified random sampling methodology that first selected residential addresses across the US, then one adult within each household. The residential addresses were grouped into high-and low-minority strata to ensure adequate representation of racial and ethnic minorities.

### Study Sample

Data were obtained from 3,865 adult respondents. The overall household response rate was 30.3%. All H5C3 responses were weighted to reflect selection probabilities adjust for non-response to provide a nationally representative sample with regard to age, gender, education, race, ethnicity, and census region. Additional details about H5C3 may be found elsewhere.

### Study Measures

Clinical trial knowledge was measured using a three-point Likert scale (no knowledge, a little knowledge, and a lot of knowledge). Ever being invited to participation in a clinical trial (yes/no) and participating in a clinical trial (yes/no among those invited) were combined into a clinical trial participation variable with three categories (never invited, invited but did not participate, invited and participated). Binary indicators of digital technology use in the past year included looking for health or medical information, communicating with a doctor’s office, looking up medical test results, and making medical appointments. Binary indicators of digital health engagement in the past year included sharing health information on social media, participating in a health forum or support group, watching health-related videos on YouTube, and awareness of ClinicalTrials.gov.

Demographics included age (18-34, 35-49, 50-64, 65-74, 75+), gender (male, female), sexual orientation (heterosexual, gay or lesbian, bisexual, other), race and ethnicity (non-Hispanic [NH] White, NH Black, Hispanic, NH Asian, NH Other), education (less than high school, high school diploma, some college, college graduate or higher), and urban-rural status (metropolitan, non-metro). Health factors included health insurance status (yes, no), lifetime chronic diseases (none, one or more), healthcare provider access (yes, no), and general health (excellent, very good, good, fair, poor).

### Statistical Analyses

Our initial analyses examined weighted frequencies for the dependent and independent variables. We then conducted cross-tabulations to examine bivariate differences in the prevalence of clinical trial knowledge and participation across the digital health variables as well as demographic and health correlates. To test our primary hypothesis, we conducted ordinal regression to evaluate the relationship between clinical trial knowledge with health technology use and digital health engagement. This calculated adjust rate ratios (aRR) and 95% confidence intervals (CI) for the health technology use and digital health engagement variables associated with clinical trial knowledge. To test the second hypothesis, we fit a multinomial logistic regression model to examine multivariate associations between clinical trial participation with health technology use and digital health engagement. This yielded aRR’s and 95% CI’s for the health technology use and digital health engagement variables associated with clinical trial participation. All analyses incorporated the overall sample weight and post-stratification variables to account for the complex sampling methods of H5C4. Multiple imputation with chained equations was used to account for missing data. A two-sided p-value of 0.05 denoted statistical significance. R Studio version 4.3.1 was used to execute all analyses, using packages *mice* and *svyVGAM*.^23,24^

### Results

The weighted proportions of the digital health variables, demographics, and health correlates are shown in Table 1. Overall, the average age was 48.4 years (SD=18.3) and 50.8% of participants were female. Most of the sample was NH White (63.2%), heterosexual (92.6%), from a metropolitan area (87.8%), and had health insurance (90.9%).

**Table 1.** Overall distribution of demographic and health correlates.

| Characteristics | Raw N=3,865 |
| --- | --- |
|  | Weighted % (95% CI) |
| Age |  |
| 18-34 | 27.1 (25.3-28.9) |
| 35-49 | 24.6 (22.5-26.7) |
| 50-64 | 27.4 (25.6-29.2) |
| 65-74 | 12.3 (11.8-12.8) |
| 75+ | 8.7 (8.2-9.1) |
| Gender |  |
| Male | 49.2 (48.3-50.2) |
| Female | 50.8 (49.8-51.7) |
| Sexual Orientation |  |
| Heterosexual | 92.6 (90.9-94.2) |
| Gay or lesbian | 2.9 (1.8-4.3) |
| Bisexual | 2.8 (1.9-3.8) |
| Other | 1.7 (1.0-2.5) |
| Race and Ethnicity |  |
| NH White | 63.2 (62.4-64.0) |
| NH Black | 11.3 (10.5-12.2) |
| Hispanic | 16.7 (16.2-17.2) |
| NH Asian | 5.6 (4.8-6.4) |
| NH Other | 3.2 (2.7-3.7) |
| Educational Attainment |  |
| Less than high school | 8.0 (6.4-9.7) |
| High school diploma | 22.4 (20.6-24.3) |
| Some college | 38.8 (37.1-40.6) |
| College graduate or higher | 30.7 (30.0-31.5) |
| Urban-Rural Status |  |
| Metropolitan | 87.8 (86.2-89.2) |
| Non-metropolitan | 12.2 (10.8-13.8) |
| Health Insurance |  |
| Yes | 90.9 (90.5-91.3) |
| No | 9.1 (8.7-9.5) |
| Lifetime Chronic Diseases |  |
| None | 41.3 (39.2-43.4) |
| One or more | 58.7 (56.6-60.8) |
| Regular Provider Access |  |
| No | 37.8 (35.3-40.3) |
| Yes | 62.2 (59.7-64.7) |
| General Health |  |
| Excellent | 12.3 (10.7-14.1) |
| Very good | 37.3 (34.9-39.8) |
| Good | 36.2 (34.0-38.5) |
| Fair | 12.1 (10.4-13.8) |
| Poor | 2.0 (1.5-2.6) |
| Past-Year Digital Technology Use |  |
| Looked for health/medical info | 72.5 (70.2-74.7) |
| Communicated with doctors office | 46.9 (44.6-49.2) |
| Looked up medical test results | 42.0 (39.0-45.0) |
| Made appointments with provider | 49.3 (46.6-52.0) |
| Past-Year Digital Health Engagement |  |
| Shared health info on social media | 14.1 (12.3-15.9) |
| Participated in forum or support group | 9.7 (8.3-11.3) |
| Watched health-related videos on YouTube | 40.4 (37.8-42.9) |
| Heard of ClinicalTrials.gov | 7.0 (5.6-8.6) |

The most common digital technology use in the past year was to search for health and medical information (72.5%) followed by making medical appointments (49.3%), communicating with a doctor’s office (46.9%), and looking up medical results (42.0%). For digital health engagement in the past year, watching health-related YouTube videos was the most common (40.4%) followed by sharing health information on social media platforms (14.1%), participating in a health forum or support group (9.7%), and awareness of ClinicalTrials.gov (7.0%).

Table 2 shows the overall prevalence of clinical trial knowledge and multivariate associations. Nearly half of the sample (48.9%) reported a little knowledge, 41.7% reported no knowledge and 9.5% reported a lot of knowledge. In the ordinal regression models, awareness of ClinicalTrials.gov was the largest association with clinical trial knowledge (aRR=7.60, 95% CI=4.82-12.00). Significant associations were also observed for searching for health and medical information (aRR=1.35, 95% CI=1.06-1.71), communicating with a doctor’s office (aRR=1.64, 95% CI=1.25-2.14), and watching health-related YouTube videos (aRR=1.36, 95% CI=1.02-1.82). Among the demographics, higher education (aOR=3.78, 95% CI=2.16-6.60) and provider access (aRR=1.85, 95% CI=1.22-2.22) were associated with higher clinical trial knowledge.

**Table 2.** Results from ordinal logistic regression of demographic and health correlates of clinical trial knowledge.

| Characteristics | Level of Clinical Trial Knowledge |  |  | aRR (95% CI) |
| --- | --- | --- | --- | --- |
|  | No Knowledge<br>Weighted % (95% CI) | A Little Knowledge<br>Weighted % (95% CI) | A Lot of Knowledge<br>Weighted % (95% CI) |  |
| Overall (N=3865) | 41.7 (38.9-44.4) | 48.9 (46.3-51.5) | 9.5 (7.9-11.2) | — |
| Age |  |  |  |  |
| 18-34 | 43.4 (36.3-50.6) | 45.7 (39.1-52.5) | 10.9 (7.1-15.3) | 1.00 |
| 35-49 | 37.3 (31.7-43.0) | 51.6 (45.1-58.2) | 11.1 (7.3-15.5) | 1.11 (0.74-1.65) |
| 50-64 | 40.8 (36.9-44.7) | 51.4 (47.1-55.6) | 7.9 (6.3-9.6) | 0.96 (0.66-1.39) |
| 65-74 | 36.7 (31.7-41.9) | 54.6 (49.2-59.9) | 8.7 (6.5-11.2) | 1.19 (0.75-1.89) |
| 75+ | 58.3 (51.8-64.7) | 35.0 (29.5-40.8) | 6.6 (4.2-9.5) | 0.68 (0.39-1.19) |
| Gender |  |  |  |  |
| Male | 42.6 (38.0-47.3) | 48.1 (43.6-52.6) | 9.3 (7.1-11.8) | 1.00 |
| Female | 40.7 (37.2-44.3) | 49.7 (46.2-53.2) | 9.6 (8.0-11.3) | 0.91 (0.70-1.18) |
| Sexual Orientation |  |  |  |  |
| Heterosexual | 42.3 (39.4-45.1) | 48.5 (45.8-51.3) | 9.2 (7.5-10.9) | 1.00 |
| Gay or lesbian | 24.5 (12.5-39.0) | 66.0 (52.7-78.1) | 9.5 (3.8-17.5) | 1.40 (0.48-4.05) |
| Bisexual | 36.2 (21.3-52.7) | 42.2 (26.2-59.2) | 21.5 (9.2-37.2) | 1.86 (0.88-3.95) |
| Other | 45.1 (22.1-69.4) | 49.0 (26.4-71.8) | 5.9 (2.0-11.6) | 0.73 (0.25-2.14) |
| Race and Ethnicity |  |  |  |  |
| NH White | 38.1 (35.0-41.2) | 51.9 (48.7-55.1) | 10.0 (8.0-12.3) | 1.00 |
| NH Black | 43.5 (35.4-51.7) | 48.5 (40.9-56.1) | 8.0 (4.7-12.1) | 0.92 (0.66-1.30) |
| Hispanic | 52.0 (43.5-60.5) | 40.5 (33.3-47.9) | 7.5 (4.0-11.9) | 0.77 (0.47-1.26) |
| NH Asian | 48.1 (34.6-61.9) | 38.2 (26.6-50.4) | 13.7 (7.3-21.7) | 0.70 (0.41-1.21) |
| NH Other | 40.8 (26.4-56.0) | 53.4 (38.5-68.0) | 5.8 (3.1-9.4) | 0.81 (0.47-1.40) |
| Educational Attainment |  |  |  |  |
| Less than high school | 68.1 (57.2-78.0) | 29.3 (19.5-40.2) | 2.7 (1.3-4.4) | 1.00 |
| High school diploma | 54.5 (48.6-60.4) | 42.7 (36.9-48.6) | 2.8 (1.7-4.1) | 1.56 (0.92-2.64) |
| Some college | 42.2 (37.9-46.5) | 51.0 (46.8-55.2) | 6.8 (4.5-9.6) | 2.01 (1.19-3.38) |
| College graduate or higher | 24.8 (20.9-28.8) | 55.8 (51.8-59.8) | 19.4 (15.8-23.4) | 3.78 (2.16-6.60) |
| Urban-Rural Status |  |  |  |  |
| Metropolitan | 40.8 (37.9-43.7) | 49.5 (46.8-52.2) | 9.7 (8.0-11.6) | 1.00 |
| Non-metropolitan | 47.7 (38.8-56.7) | 44.7 (36.3-53.3) | 7.6 (3.5-13.3) | 0.89 (0.57-1.39) |
| Health Insurance |  |  |  |  |
| Yes | 39.7 (36.8-42.6) | 50.3 (47.5-53.1) | 10.0 (8.3-11.9) | 1.00 |
| No | 61.1 (48.4-73.0) | 35.0 (24.0-46.9) | 4.0 (1.5-7.5) | 0.60 (0.33-1.10) |
| Lifetime Chronic Diseases |  |  |  |  |
| None | 41.6 (36.5-46.9) | 49.6 (44.6-54.7) | 8.7 (7.0-10.6) | 1.00 |
| One or more | 41.7 (38.0-45.4) | 48.4 (44.9-51.8) | 10.0 (8.0-12.2) | 1.00 (0.73-1.37) |
| Provider Access |  |  |  |  |
| No | 51.5 (46.8-56.2) | 41.1 (36.6-45.7) | 7.4 (5.5-9.6) | 1.00 |
| Yes | 35.7 (32.2-39.2) | 53.6 (50.1-57.1) | 10.7 (8.6-13.0) | 1.64 (1.22-2.22) |
| General Health |  |  |  |  |
| Excellent | 32.8 (24.5-41.6) | 49.3 (42.2-56.4) | 18.0 (12.5-24.2) | 1.55 (0.63-3.85) |
| Very good | 39.8 (35.2-44.5) | 50.1 (45.7-54.5) | 10.1 (7.8-12.7) | 1.16 (0.53-2.55) |
| Good | 43.4 (38.9-48.0) | 50.2 (45.3-55.0) | 6.4 (4.6-8.5) | 1.05 (0.47-2.39) |
| Fair | 48.7 (40.8-56.6) | 43.3 (36.0-50.7) | 8.0 (4.5-12.5) | 1.05 (0.43-2.54) |
| Poor | 56.0 (38.3-73.0) | 34.9 (18.4-53.6) | 9.0 (3.8-16.1) | 1.00 |
| Past-Year Digital Technology Use |  |  |  |  |
| Looked for health/medical info | 35.6 (32.6-38.7) | 52.8 (49.8-55.8) | 11.6 (9.5-13.8) | 1.35 (1.06-1.71) |
| Communicated with doctors office | 29.3 (26.0-32.8) | 56.6 (53.1-60.1) | 14.0 (11.2-17.1) | 1.64 (1.25-2.14) |
| Looked up medical test results | 31.6 (27.4-36.0) | 54.0 (50.5-57.4) | 14.4 (11.5-17.6) | 0.94 (0.67-1.33) |
| Made appointments with provider | 34.7 (31.1-38.4) | 51.4 (48.1-54.8) | 13.9 (11.3-16.7) | 1.07 (0.82-1.39) |
| Past-Year Digital Health Engagement |  |  |  |  |
| Shared health info on social media | 28.6 (23.5-33.9) | 58.3 (52.1-64.4) | 13.1 (9.4-17.3) | 1.17 (0.87-1.58) |
| Participated in forum or support group | 19.3 (13.8-25.5) | 64.5 (55.6-72.8) | 16.2 (9.6-24.2) | 1.20 (0.84-1.72) |
| Watched health-related videos on YouTube | 33.2 (29.1-37.4) | 53.2 (48.7-57.6) | 13.6 (10.9-16.6) | 1.36 (1.02-1.82) |
| Heard of clinicaltrials.gov | 10.0 (6.6-14.1) | 46.8 (34.7-59.0) | 43.2 (31.5-55.3) | 7.60 (4.82-12.00) |

Table 3 shows the prevalence of clinical trial participation and multivariate associations. Overall, 9.7% of respondents were invited to participate in a clinical trial, with 5.3% who did not participate and 4.4% who participated. Digital health variables of making medical appointments (aRR=1.79, 95% CI=1.07-2.99) and awareness of ClinicalTrials.gov (aRR=2.60, 95% CI=1.23-5.54) were significantly associated with participation in the multinomial logistic regression model. Higher education was also associated with an increase in participation (aRR=5.46, 95% CI=1.82-16.38). Participants aged 65-74 years (aRR=2.36, 95% CI=1.00-5.58) and 75 years and older (aRR=2.93, 95% CI=1.17-7.37) were more likely to participate than those aged 18-34 years. Among those who participated in clinical trials, participants were less likely to be NH Asian compared to NH White (aRR=0.14, 95% CI=0.03-0.61). For those who were invited and did not participate, those participating in health forums or support groups (aRR=2.32, 95% CI=1.22-4.39) were more likely to have not participated. Participants 50-64 (aRR=2.37, 95% CI=1.05-5.32) and 65-74 (aRR=2.98, 95% CI=1.28-6.90) years old were more likely than 18–34-year-olds to have not participated. NH Blacks were more likely than NH Whites to have not participated (aRR=2.25, 95% CI=1.23-4.11).

**Table 3.** Results from multinomial logistic regression of demographic and health correlates of clinical trial participation.

| Characteristics | Not Invited | Invited, Did not Participate (n=231) |  | Invited and Participated (n=196) |  |
| --- | --- | --- | --- | --- | --- |
|  | % (95% CI) | % (95% CI) | aRR (95% CI) | % (95% CI) | aRR (95% CI) |
| Overall (N=3865) | 90.3 (88.5-92.0) | 5.3 (3.9-6.8) | — | 4.4 (3.3-5.7) | — |
| Age |  |  |  |  |  |
| 18-34 | 94.2 (91.4-96.5) | 2.4 (1.0-4.2) | 1.00 | 3.4 (1.4-6.3) | 1.00 |
| 35-49 | 89.9 (86.4-92.9) | 5.9 (3.4-9.0) | 2.45 (0.99-6.04) | 4.2 (2.3-6.7) | 1.37 (0.54-3.47) |
| 50-64 | 88.7 (84.6-92.2) | 6.7 (3.8-10.3) | 2.37 (1.05-5.32) | 4.6 (3.1-6.5) | 1.63 (0.76-3.51) |
| 65-74 | 86.2 (82.9-89.3) | 8.1 (5.6-11.0) | 2.98 (1.28-6.90) | 5.7 (3.8-7.9) | 2.36 (1.00-5.58) |
| 75+ | 90.3 (86.4-93.6) | 4.1 (2.4-6.2) | 1.52 (0.82-2.80) | 5.6 (3.4-8.4) | 2.93 (1.17-7.37) |
| Gender |  |  |  |  |  |
| Male | 90.8 (87.8-93.4) | 5.2 (3.1-7.8) | 1.00 | 4.1 (2.6-5.9) | 1.00 |
| Female | 89.9 (87.8-91.8) | 5.4 (4.2-6.6) | 0.87 (0.52-1.45) | 4.8 (3.2-6.6) | 1.00 (0.54-1.86) |
| Sexual Orientation |  |  |  |  |  |
| Heterosexual | 90.8 (89.0-92.4) | 5.0 (3.8-6.5) | 1.00 | 4.2 (3.1-5.4) | 1.00 |
| Gay or lesbian | 78.2 (58.9-92.6) | 12.6 (2.6-28.5) | 2.48 (0.43-14.50) | 9.2 (0.6-26.1) | 2.24 (0.23-21.44) |
| Bisexual | 85.1 (73.4-93.9) | 5.4 (1.3-11.9) | 1.34 (0.45-3.96) | 9.5 (2.8-19.6) | 2.2 (0.65-7.42) |
| Other | 94.2 (83.7-99.5) | 4.8 (0.1-16.0) | 0.82 (0.03-23.92) | 1.0 (0.1-2.8) | 0.2 (0.04-1.11) |
| Race and Ethnicity |  |  |  |  |  |
| NH White | 90.6 (88.3-92.7) | 4.8 (3.2-6.7) | 1.00 | 4.6 (3.4-6.0) | 1.00 |
| NH Black | 84.4 (79.4-88.9) | 11.2 (7.3-15.9) | 2.25 (1.23-4.11) | 4.3 (2.0-7.4) | 1.12 (0.50-2.51) |
| Hispanic | 91.7 (87.2-95.3) | 3.6 (1.6-6.4) | 1.13 (0.48-2.66) | 4.6 (1.9-8.4) | 1.43 (0.62-3.31) |
| NH Asian | 96.4 (92.4-99.0) | 2.9 (0.7-6.4) | 0.61 (0.17-2.19) | 0.7 (0.1-1.8) | 0.14 (0.03-0.61) |
| NH Other | 87.3 (77.5-94.6) | 6.1 (2.3-11.6) | 1.33 (0.49-3.60) | 6.5 (1.0-16.4) | 1.46 (0.37-5.74) |
| Educational Attainment |  |  |  |  |  |
| Less than high school | 94.6 (91.4-97.1) | 4.3 (2.1-7.2) | 1.00 | 1.2 (0.4-2.4) | 1.00 |
| High school diploma | 91.6 (87.6-94.8) | 5.1 (2.5-8.5) | 1.11 (0.47-2.65) | 3.4 (1.7-5.6) | 2.86 (0.89-9.24) |
| Some college | 90.8 (88.3-92.9) | 5.4 (3.6-7.5) | 1.08 (0.39-2.99) | 3.9 (2.4-5.6) | 3.00 (1.03-8.77) |
| College graduate or higher | 87.7 (84.8-90.4) | 5.6 (3.6-7.9) | 1.20 (0.41-3.52) | 6.7 (4.7-9.1) | 5.46 (1.82-16.38) |
| Urban-Rural Status |  |  |  |  |  |
| Metropolitan | 89.9 (88.1-91.6) | 5.6 (4.3-7.1) | 1.00 | 4.5 (3.4-5.7) | 1.00 |
| Non-metropolitan | 93.1 (88.0-96.9) | 3.0 (0.6-7.1) | 0.47 (0.14-1.65) | 3.9 (1.3-7.8) | 0.91 (0.36-2.31) |
| Health Insurance |  |  |  |  |  |
| Yes | 89.9 (87.9-91.7) | 5.7 (4.2-7.3) | 1.00 | 4.4 (3.3-5.7) | 1.00 |
| No | 94.5 (87.9-98.7) | 1.1 (0.2-2.8) | 0.33 (0.07-1.53) | 4.3 (0.6-11.3) | 1.65 (0.27-10.03) |
| Lifetime Chronic Diseases |  |  |  |  |  |
| None | 94.7 (93.0-96.2) | 2.1 (1.3-3.0) | 1.00 | 3.2 (1.9-4.8) | 1.00 |
| One or more | 87.2 (84.6-89.7) | 7.5 (5.4-9.9) | 3.18 (1.81-5.62) | 5.3 (3.8-7.0) | 1.55 (0.88-2.72) |
| Provider Access |  |  |  |  |  |
| No | 94.2 (91.9-96.1) | 2.6 (1.5-4.1) | 1.00 | 3.2 (1.8-5.0) | 1.00 |
| Yes | 88.0 (85.3-90.4) | 6.9 (4.9-9.2) | 2.01 (1.02-3.95) | 5.2 (3.8-6.7) | 1.29 (0.73-2.28) |
| General Health |  |  |  |  |  |
| Excellent | 91.3 (86.2-95.2) | 4.6 (1.8-8.7) | 1.57 (0.42-5.91) | 4.1 (1.4-8.1) | 0.35 (0.04-2.74) |
| Very good | 90.9 (88.1-93.4) | 4.8 (3.0-6.9) | 1.11 (0.47-2.62) | 4.3 (2.8-6.2) | 0.33 (0.06-1.79) |
| Good | 91.2 (88.7-93.4) | 4.9 (3.3-6.8) | 0.88 (0.36-2.14) | 3.9 (2.4-5.7) | 0.28 (0.04-1.95) |
| Fair | 86.1 (80.6-90.8) | 8.3 (4.9-12.5) | 1.30 (0.44-3.87) | 5.6 (2.6-9.5) | 0.42 (0.06-3.16) |
| Poor | 82.2 (67.4-93.2) | 6.7 (2.8-12.0) | 1.00 | 11.2 (1.6-27.7) | 1.00 |
| Past-Year Digital Technology Use |  |  |  |  |  |
| Looked for health/medical info | 89.5 (87.5-91.4) | 5.3 (4.0-6.8) | 0.87 (0.47-1.60) | 5.1 (3.8-6.7) | 1.55 (0.84-2.87) |
| Communicated with doctors office | 88.5 (86.2-90.7) | 6.0 (4.3-7.9) | 0.78 (0.42-1.45) | 5.5 (3.8-7.6) | 0.83 (0.48-1.42) |
| Looked up medical test results | 88.2 (85.4-90.7) | 6.1 (4.2-8.2) | 0.83 (0.44-1.58) | 5.8 (3.8-8.1) | 0.99 (0.49-2.03) |
| Made appointments with provider | 87.7 (84.8-90.3) | 6.4 (4.3-8.8) | 1.32 (0.80-2.17) | 5.9 (4.1-8.1) | 1.79 (1.07-2.99) |
| Past-Year Digital Health Engagement |  |  |  |  |  |
| Shared health info on social media | 85.0 (79.4-89.7) | 9.4 (5.7-13.9) | 1.45 (0.80-2.64) | 5.7 (2.6-9.7) | 0.86 (0.35-2.1) |
| Participated in forum or support group | 77.9 (71.2-84.0) | 12.7 (7.8-18.6) | 2.32 (1.22-4.39) | 9.3 (5.1-14.6) | 1.88 (0.91-3.87) |
| Watched health-related videos on YouTube | 88.0 (84.7-91.0) | 6.7 (4.4-9.4) | 1.36 (0.69-2.68) | 5.3 (3.6-7.4) | 1.21 (0.68-2.16) |
| Heard of ClinicalTrials.gov | 80.0 (72.7-86.4) | 8.2 (4.9-12.2) | 1.48 (0.77-2.83) | 11.8 (5.8-19.6) | 2.60 (1.23-5.54) |

## Discussion

This study evaluated the association between health technology use and digital health engagement with clinical trial knowledge and participation in a general population sample. The main results partially supported our primary hypothesis, as searching for health and medical information, communicating with a doctor’s office, watching health-related videos on YouTube, and awareness of ClinicalTrials.gov were significantly associated with higher clinical trial knowledge. The secondary hypothesis was also partially supported, with making appointments with medical providers and awareness of ClinicalTrials.gov being significantly associated with clinical trial participation. Awareness of ClinicalTrials.gov was the strongest correlate for both knowledge and participation after adjusting for demographics and health factors, suggesting that promotion of digital research platforms should be a component of patient and provider education programs focused on increasing clinical trial participation. The current study builds upon previous research on clinical trial knowledge and participation by demonstrating important ways that a variety of digital technologies and platforms may help to increase inclusion in research.^14–21,25,26^

We found that multiple measures of digital technology use to engage with providers and electronic health platforms were associated with higher clinical trial awareness and participation. Because the majority of clinical research participants report learning about studies from their healthcare provider,^27,28^ promoting uptake of digital technologies may facilitate additional opportunities for provider interactions that increase awareness about the option of clinical research participation.^29,30^ For example, emergent research has found evidence that integrating clinical research procedures into routine clinical care workflows within electronic health record systems—both patient– and provider-facing—can boost recruitment rates and streamline enrollment procedures.^31,32^ Other studies have demonstrated the acceptability of clinical research decision-making tools and artificial intelligence platforms in optimizing key research processes.^33–37^

Nevertheless, among the 10% who received an invitation to participate in a clinical trial, less than half went on to participate. In particular, Black respondents were relatively more likely to have been invited but not participated. This may be a function of more severe patterns of comorbidities and adverse SDOH among Black populations that increase the likelihood of service utilization and being invited to participate in clinical trials.^38,39^ Similarly, other HINTS studies have found that Black respondents are less likely to report being influenced by doctors and family members to participate in clinical trials,^12^ which is also consistent with longstanding distrust of the medical community that has been exacerbated by the COVID-19 pandemic.^40,41^ In addition, Asians had the lowest rates of being invited (3.6% vs 9.7% overall) and were 86% less likely to have participated. As provider access and communications were associated with both clinical trial knowledge and participation, the effects observed in this study may be related to cultural factors (e.g., social stigma, familial shame) shown to influence low rates of service utilization among Asians compared to other racial groups.^42^ Moreover, recent studies indicate that Asians as an aggregate category appear to have more favorable quality of life than NH Whites, masking significant variability in morbidity and mortality among Asian subpopulations.^43–45^ This suggests that certain Asian subpopulations may be less likely to receive medical care, thereby decreasing opportunities during medical encounters and follow-ups to be invited into a clinical trial.^11,46,47^ Additional research is needed to evaluate the impact of disaggregating race and ethnicity on differences in clinical trial knowledge and participation,^43,48^ including implications for recent changes by the US Office of Management and Budget in how race and ethnicity will be collected to reduce misclassification.

Several limitations are acknowledged. The cross-sectional design precludes the ability to infer temporality between the significant correlates of clinical trial knowledge and participation. Pragmatic trials and novel observational studies (e.g., natural experiments, regression discontinuity, observational-implementation hybrid approach) will be necessary to more rigorously evaluate the real-world effectiveness of digital health interventions to increase clinical trial participation.^49–53^ While this study employed data generalizable to the adult population, the small proportion of clinical trial participation reduced the precision of estimates in certain demographic subpopulations. The limited sample size may have also led to Type II errors for race, ethnicity, and sexual orientation. Upcoming HINTS cycles should re-administer the clinical trials questions to allow for larger analytical samples and more rigorous investigation across subpopulations. Similarly, access to digital technologies, availability of broadband internet, and digital health literacy have been found to be lower in a variety of health disparity populations, yet significant gaps remain about the individual and structural factors driving digital health inequities.^18,19,54^ Intersectionality and participatory action approaches will be necessary to better align research participation efforts with the needs of specific subpopulations.^55–58^ In addition, future studies should focus on developing strategies to counter mis– and dis-information rapidly propagated on social media and other digital platforms that increase science rejection.^59–61^ Exposure to such information may contribute to misperceptions about the benefits of clinical research that conflict with established scientific evidence and reduce the likelihood of clinical trial participation.^62^

Based upon the study’s findings, large-scale health promotion campaigns should be implemented to distribute digital resources about clinical trial participation. Further adaptations should be considered based upon demographic and health factors. Intervention studies should determine the real-world effectiveness of integrating clinical research procedures into routine clinical care workflows and electronic health record systems as well as testing other emergent health technologies like AI. Taken together, the findings from this study will inform the development of quality improvement programs that improve clinical trial participation.

## Data Availability

All data produced in the present study are available upon reasonable request to the authors.
The Heath Information National Trends Survey dataset is available at https://hints.cancer.gov/.

## Acknowledgments

This study was supported by an unrestricted grant through the Abbott Laboratories Diversity in Clinical Research Initiative. Dr. Peiper reports research support through the National Institutes of Health and Centers for Disease Control and Prevention (CDC). Dr. Peiper reports scientific advisory fees and stock options from Meru Health. Mr. Brown reports research support through the CDA Foundation and scientific consultation fees from Meru Health.

