## Supplementary Tables for "Effects of Health Technology Use and Digital Health Engagement on Clinical Trial Participation: Findings from the Health Information National Trends Survey"

Supplemental Table 1. Overall distribution of demographic and health correlates

| Characteristics | Raw N=2,814<br>Weighted (95% CI) |
|---|---|
| Age | |
| 18-34 | 26.4 (24.4-28.4) |
| 35-49 | 25.0 (22.7-27.3) |
| 50-64 | 28.0 (25.9-30.2) |
| 65-74 | 11.9 (11.4-12.5) |
| 75+ | 8.7 (8.0-9.4) |
| Gender | |
| Male | 48.9 (47.8-49.9) |
| Female | 51.1 (50.1-52.2) |
| Sexual Orientation | |
| Heterosexual | 92.6 (90.8-94.2) |
| Gay or lesbian | 2.8 (1.9-4.0) |
| Bisexual | 2.8 (1.7-4.2) |
| Other | 1.8 (1.1-2.6) |
| Race and Ethnicity | |
| NH White | 62.1 (61.0-63.1) |
| NH Black | 11.4 (10.8-12.0) |
| Hispanic | 16.8 (16.4-17.2) |
| NH Asian | 5.6 (4.8-6.5) |
| NH Other | 4.1 (3.3-5.0) |
| Educational Attainment | |
| Less than high school | 8.1 (6.6-9.8) |
| High school diploma | 22.3 (20.6-24.1) |
| Some college | 39.1 (37.3-41.0) |
| College graduate or higher | 30.4 (29.7-31.2) |
| Urban-Rural Status | |
| Metropolitan | 87.8 (86.2-89.2) |
| Non-metropolitan | 12.2 (10.8-13.8) |
| Health Insurance | |
| Yes | 90.8 (90.4-91.3) |
| No | 9.2 (8.7-9.6) |
| Lifetime Chronic Diseases | |
| None | 41.3 (39.2-43.4) |
| One or more | 58.7 (56.6-60.8) |
| Regular Provider Access | |
| No | 37.8 (35.4-40.3) |
| Yes | 62.2 (59.7-64.6) |
| General Health | |
| Excellent | 12.3 (10.7-14.1) |
| Very good | 37.5 (35.1-39.9) |
| Good | 36.1 (33.9-38.4) |
| Fair | 12.0 (10.4-13.8) |
| Poor | 2.0 (1.5-2.6) |
| Past-Year Digital Technology Use | |
| Looked for health/medical info | 72.6 (70.4-74.7) |
| Communicated with doctors office | 47.0 (44.7-49.3) |
| Looked up medical test results | 42.0 (39.0-45.1) |
| Made appointments with provider | 49.3 (46.5-52.1) |
| Past-Year Digital Health Engagement | |
| Shared health info on social media | 14.4 (12.7-16.1) |
| Participated in forum or support group | 9.8 (8.4-11.3) |
| Watched health-related videos on YouTube | 40.8 (38.3-43.3) |
| Heard of ClinicalTrials.gov | 7.0 (5.6-8.6) |

Supplemental Table 2. Results from multinomial logistic regression of demographic and health correlates of clinical trial participation

| Characteristics | Not Invited | Invited, Did not Participate (n=231) | | Invited and Participated (n=196) | |
|---|---|---|---|---|---|
| | (95% CI) | (95% CI) | aRR (95% CI) | (95% CI) | aRR (95% CI) |
| Overall (N=2,814) | 90.2 (88.3-91.9) | 5.2 (4.0-6.6) | — | 4.6 (3.4-6.0) | — |
| Age | | | | | |
| 18-34 | 94.3 (90.9-96.9) | 2.3 (0.9-4.5) | 1.00 | 3.4 (1.2-6.6) | 1.00 |
| 35-49 | 91.3 (87.7-94.3) | 4.2 (2.3-6.7) | 1.73 (0.59-5.00) | 4.5 (2.2-7.5) | 1.24 (0.52-2.94) |
| 50-64 | 89.5 (85.5-92.9) | 5.9 (3.2-9.5) | 1.96 (0.75-5.13) | 4.6 (2.9-6.6) | 1.15 (0.61-2.17) |
| 65-74 | 84.4 (80.1-88.3) | 9.2 (6.0-12.9) | 2.70 (0.91-7.99) | 6.4 (4.1-12.9) | 1.73 (0.75-3.95) |
| 75+ | 89.2 (83.1-94.1) | 5.0 (2.3-8.7) | 2.52 (0.72-8.84) | 5.7 (2.4-10.3) | 1.84 (0.79-4.28) |
| Gender | | | | | |
| Male | 90.8 (87.7-93.5) | 4.9 (2.8-7.5) | 1.00 | 4.3 (2.6-6.5) | 1.00 |
| Female | 90.8 (88.4-92.9) | 4.8 (3.5-5.8) | 0.83 (0.52-1.33) | 4.6 (3.9-6.8) | 0.93 (0.53-1.63) |
| Sexual Orientation | | | | | |
| Heterosexual | 91.1 (89.3-92.7) | 4.6 (3.3-6.1) | 1.00 | 4.3 (3.3-6.1) | 1.00 |
| Gay or lesbian | 78.0 (53.7-94.9) | 9.1 (3.0-18.1) | 2.35 (0.54-10.25) | 12.9 (0.2-44.2) | 3.55 (0.52-24.39) |
| Bisexual | 91.4 (82.1-97.6) | 4.2 (1.0-9.4) | 1.47 (0.51-4.29) | 4.4 (0.7-11.0) | 1.62 (0.41-6.34) |
| Other | 92.4 (71.4-99.9) | 7.1 (0.1-28.4) | 0.73 (0.03-20.97) | 0.5 (0.1-2.3) | 0.58 (0.08-4.19) |
| Race and Ethnicity | | | | | |
| NH White | 90.8 (88.7-92.7) | 4.3 (2.9-6.0) | 1.00 | 4.8 (3.6-6.2) | 1.00 |
| NH Black | 82.6 (76.1-88.2) | 12.5 (7.6-18.4) | 2.36 (1.36-4.12) | 4.9 (1.8-9.5) | 1.01 (0.48-2.14) |
| Hispanic | 94.9 (90-98.2) | 1.6 (0.7-3.0) | 0.91 (0.42-1.96) | 3.5 (0.8-7.9) | 1.53 (0.73-3.21) |
| NH Asian | 97.5 (94-99.5) | 2.1 (0.3-5.6) | 0.29 (0.05-1.70) | 0.4 (0.1-1.5) | 0.05 (0.00-3895.59) |
| NH Other | 83.7 (70.3-93.7) | 8.2 (2.9-16.1) | 1.55 (0.72-3.33) | 8 (0.8-21.7) | 1.51 (0.45-5.07) |
| Educational Attainment | | | | | |
| Less than high school | 94.3 (89.3-97.8) | 4.8 (1.6-9.6) | 1.00 | 0.9 (0.1-2.3) | 1.00 |
| High school diploma | 93.7 (89.5-96.9) | 3.8 (1.2-7.9) | 0.87 (0.40-1.91) | 2.5 (0.9-4.8) | 3.30 (0.80-13.66) |
| Some college | 92.0 (89.6-94.1) | 4.2 (2.8-5.9) | 1.05 (0.42-2.66) | 3.7 (2.2-5.7) | 2.69 (0.97-7.51) |
| College graduate or higher | 86.7 (83.4-89.8) | 5.9 (3.9-8.4) | 1.16 (0.47-2.85) | 7.3 (5.0-10.1) | 5.42 (1.57-18.71) |
| Urban-Rural Status | | | | | |
| Metropolitan | 90.4 (88.6-92.1) | 5.1 (3.8-6.6) | 1.00 | 4.4 (3.3-5.7) | 1.00 |
| Non-metropolitan | 93.4 (87.8-97.4) | 1.6 (0.2-4.1) | 0.22 (0.05-0.88) | 5.0 (1.5-10.2) | 1.31 (0.57-2.99) |
| Health Insurance | | | | | |
| Yes | 90.4 (88.5-92.1) | 5.1 (3.7-6.6) | 1.00 | 4.5 (3.4-5.8) | 1.00 |
| No | 94.9 (86.6-99.4) | 1.2 (0.1-3.4) | 0.3 (0.07-1.38) | 3.9 (0.1-12.5) | 1.58 (0.27-9.29) |
| Lifetime Chronic Diseases | | | | | |
| None | 95.7 (94.0-97.0) | 1.7 (1.0-2.7) | 1.00 | 2.6 (1.4-4.2) | 1.00 |
| One or more | 87.1 (84.2-89.8) | 7.0 (4.9-9.4) | 2.36 (1.3-4.29) | 5.9 (4.1-8.0) | 1.96 (1.10-3.49) |
| Provider Access | | | | | |
| No | 94.7 (92.2-96.8) | 2.2 (1.0-3.9) | 1.00 | 3.0 (1.6-4.9) | 1.00 |
| Yes | 88.4 (85.8-90.9) | 6.2 (4.4-8.3) | 1.42 (0.74-2.75) | 5.4 (3.8-7.2) | 1.64 (0.90-2.99) |
| General Health | | | | | |
| Excellent | 91.7 (85.3-96.3) | 4.3 (1.1-9.4) | 1.45 (0.40-5.27) | 4.0 (1.1-8.8) | 0.30 (0.05-1.90) |
| Very good | 92.0 (89.2-94.4) | 4.1 (2.4-6.2) | 0.95 (0.37-2.44) | 3.9 (2.4-5.7) | 0.33 (0.08-1.37) |

| | | | | | |
|---|---|---|---|---|---|
| Good | 91.3 (88.8-93.5) | 4.6 (3.1-6.4) | 0.92 (0.35-2.41) | 4.1 (2.5-6.1) | 0.25 (0.05-1.26) |
| Fair | 86.5 (80.9-91.2) | 7.0 (4.0-10.8) | 1.40 (0.45-4.34) | 6.5 (3.0-11.3) | 0.36 (0.07-1.99) |
| Poor | 71.3 (49.0-89.3) | 10.6 (4.1-19.5) | 1.00 | 18.1 (1.9-45.4) | 1.00 |
| Past-Year Digital Technology Use | | | | | |
| Looked for health/medical info | 89.6 (87.4-91.5) | 5.0 (3.7-6.6) | 1.23 (0.68-2.25) | 5.4 (3.9-7.1) | 1.23 (0.55-2.73) |
| Communicated with doctors office | 88.4 (85.7-90.8) | 5.5 (3.7-7.6) | 0.75 (0.41-1.36) | 6.1 (4.0-8.6) | 0.81 (0.45-1.45) |
| Looked up medical test results | 87.7 (84.5-90.5) | 6.0 (4.1-8.1) | 0.91 (0.49-1.69) | 6.4 (4.1-9.2) | 0.96 (0.48-1.92) |
| Made appointments with provider | 88.4 (85.6-90.9) | 5.4 (3.6-7.5) | 1.49 (0.87-2.57) | 6.2 (4.2-8.7) | 1.61 (0.94-2.75) |
| Past-Year Digital Health Engagement | | | | | |
| Shared health info on social media | 85.6 (79.6-90.6) | 8.0 (4.3-12.6) | 1.52 (0.78-2.97) | 6.5 (2.9-11.3) | 1.10 (0.48-2.52) |
| Participated in forum or support group | 79.6 (72.5-85.9) | 10.2 (5.5-16.1) | 1.67 (0.75-3.72) | 10.2 (5.3-16.5) | 1.68 (0.82-3.45) |
| Watched health-related videos on YouTube | 89.2 (86.1-92.0) | 5.5 (3.5-7.8) | 1.30 (0.68-2.51) | 5.3 (3.4-7.5) | 1.33 (0.71-2.51) |
| Heard of ClinicalTrials.gov | 78.7 (69.1-87.0) | 7.3 (4.2-11.1) | 1.54 (0.77-3.08) | 14.0 (6.5-23.7) | 2.58 (1.17-5.68) |

Supplemental Table 3. Results from ordinal logistic regression of demographic and health correlates of clinical trial knowledge

| | Level of Clinical Trial Knowledge | | | |
|---|---|---|---|---|
| | No Knowledge | A Little Knowledge | A Lot of Knowledge | |
| Characteristics | Weighted (95% CI) | Weighted (95% CI) | Weighted (95% CI) | aRR (95% CI) |
| Overall (N=2,814) | 41.4 (38.7-44.2) | 49.1 (46.4-51.7) | 9.5 (8.0-11.2) | — |
| Age | | | | |
| 18-34 | 43.2 (35.2-51.3) | 45.0 (37.7-52.4) | 11.8 (7.5-17.0) | 1.00 |
| 35-49 | 35.6 (29.5-42.0) | 52.6 (45.0-60.1) | 11.8 (7.9-16.4) | 1.08 (0.71-1.64) |
| 50-64 | 37.8 (33.4-42.3) | 54.1 (49.1-59.1) | 8.1 (6.3-10.1) | 0.90 (0.63-1.30) |
| 65-74 | 30.1 (24.3-36.3) | 61.5 (55.0-67.8) | 8.3 (5.5-11.7) | 1.13 (0.72-1.78) |
| 75+ | 49.5 (41.3-57.7) | 43.2 (35.6-51.1) | 7.2 (3.7-11.8) | 0.63 (0.37-1.08) |
| Gender | | | | |
| Male | 40.7 (35.5-46.1) | 49.3 (44.2-54.4) | 10.0 (7.2-13.1) | 1.00 |
| Female | 36.6 (32.6-40.7) | 53.1 (49.0-57.2) | 10.3 (8.5-12.2) | 0.92 (0.7-1.21) |
| Sexual Orientation | | | | |
| Heterosexual | 39.5 (36.2-42.9) | 50.8 (47.5-54.0) | 9.7 (7.8-11.7) | 1.00 |
| Gay or lesbian | 16.5 (7.5-28.0) | 70.3 (52.8-85.2) | 13.2 (5.0-24.6) | 1.35 (0.44-4.16) |
| Bisexual | 31.7 (14.3-52.3) | 45.0 (24.8-66.0) | 23.4 (8.8-42.3) | 1.95 (0.95-4.04) |
| Other | 30.9 (4.7-67.1) | 63.3 (30.9-90.1) | 5.8 (0.6-16.0) | 0.78 (0.26-2.33) |
| Race and Ethnicity | | | | |
| NH White | 34.7 (31.3-38.3) | 54.4 (50.7-58.2) | 10.8 (8.6-13.3) | 1.00 |
| NH Black | 37.8 (29.1-47.0) | 54.4 (46.0-62.8) | 7.8 (3.7-13.1) | 1.02 (0.73-1.43) |
| Hispanic | 51.6 (41.2-61.8) | 40.7 (31.6-50.0) | 7.8 (3.4-13.7) | 0.74 (0.44-1.22) |
| NH Asian | 50.8 (37.5-64.0) | 33.7 (23.8-44.3) | 15.6 (6.9-26.9) | 0.52 (0.31-0.87) |
| NH Other | 38.3 (21.5-56.7) | 56.1 (38.7-72.7) | 5.6 (2.3-10.3) | 0.76 (0.45-1.26) |
| Educational Attainment | | | | |
| Less than high school | 68.4 (54.4-80.8) | 29.0 (17.0-42.7) | 2.6 (1.0-5.0) | 1.00 |
| High school diploma | 51.8 (44.3-59.4) | 46.2 (38.7-53.9) | 1.9 (1.0-3.2) | 1.60 (0.93-2.76) |
| Some college | 40.8 (35.6-46.2) | 52.8 (47.3-58.3) | 6.4 (3.7-9.7) | 2.05 (1.18-3.56) |
| College graduate or higher | 21.2 (17.1-25.7) | 56.5 (52.0-61.0) | 21.2 (17.1-25.7) | 3.93 (2.21-7.00) |
| Urban-Rural Status | | | | |
| Metropolitan | 38.1 (34.8-41.4) | 51.6 (48.5-54.7) | 10.3 (8.3-12.5) | 1.00 |
| Non-metropolitan | 43.1 (32.1-54.4) | 48.3 (37.9-58.8) | 8.6 (3.6-15.4) | 0.92 (0.59-1.42) |
| Health Insurance | | | | |
| Yes | 36.3 (32.8-39.9) | 53.0 (49.4-56.6) | 10.6 (8.7-12.8) | 1.00 |
| No | 63.5 (48.3-77.5) | 31.9 (19.5-45.9) | 4.5 (1.5-9.0) | 0.58 (0.32-1.04) |
| Lifetime Chronic Diseases | | | | |
| None | 40.2 (34.0-46.6) | 50.5 (44.4-56.6) | 9.2 (7.2-11.5) | 1.00 |
| One or more | 37.5 (33.0-42.1) | 51.7 (47.4-56.1) | 10.8 (8.4-13.4) | 0.97 (0.7-1.33) |
| Provider Access | | | | |
| No | 49.9 (44.9-54.9) | 42.7 (37.9-47.5) | 7.5 (5.3-10.0) | 1.00 |
| Yes | 32.0 (28.0-36.2) | 56.3 (51.9-60.6) | 11.7 (9.3-14.4) | 1.65 (1.23-2.2) |
| General Health | | | | |
| Excellent | 30.3 (21.6-30.9) | 49.7 (42.0-57.3) | 20.0 (13.7-27.2) | 1.56 (0.64-3.79) |
| Very good | 36.6 (31.3-47.2) | 52.0 (47.6-57.5) | 10.8 (8.2-13.7) | 1.17 (0.54-2.54) |

| | | | | |
|---|---|---|---|---|
| Good | 41.6 (36.2-47.2) | 52.0 (46.5-57.5) | 6.3 (4.3-8.7) | 1.11 (0.50-2.45) |
| Fair | 43.2 (33.5-53.1) | 48.0 (38.9-57.2) | 8.8 (4.2-14.9) | 1.06 (0.45-2.54) |
| Poor | 58.6 (40.3-75.8) | 32.6 (17.1-50.4) | 8.7 (1.9-19.9) | 1.00 |
| Past-Year Digital Technology Use | | | | |
| Looked for health/medical info | 33.1 (29.5-36.7) | 54.6 (51.0-58.2) | 12.4 (10.1-14.8) | 1.35 (1.07-1.71) |
| Communicated with doctors office | 25.8 (21.7-30.1) | 59.2 (55.1-63.3) | 15.0 (11.9-18.4) | 1.63 (1.23-2.15) |
| Looked up medical test results | 29.3 (24.4-34.4) | 55.5 (51.1-59.6) | 15.3 (12.3-18.6) | 0.92 (0.65-1.29) |
| Made appointments with provider | 31.6 (27.0-36.5) | 53.2 (49.0-57.3) | 15.2 (12.2-18.4) | 1.08 (0.84-1.41) |
| Past-Year Digital Health Engagement | | | | |
| Shared health info on social media | 25.4 (20.0-31.2) | 61.4 (54.4-68.2) | 13.2 (9.5-17.4) | 1.19 (0.90-1.59) |
| Participated in forum or support group | 16.3 (11.1-22.2) | 66.1 (57.1-74.6) | 17.6 (10.0-26.7) | 1.27 (0.89-1.80) |
| Watched health-related videos on YouTube | 32.0 (27.4-36.7) | 53.5 (48.2-58.7) | 14.6 (11.5-17.9) | 1.33 (1.00-1.77) |
| Heard of clinicaltrials.gov | 6.7 (2.7-12.4) | 42.2 (31.1-53.7) | 51.1 (39.6-62.5) | 8.48 (5.41-13.3) |
